# Subtyping First-Episode Psychosis based on Longitudinal Symptom Trajectories Using Machine Learning

**DOI:** 10.1101/2024.09.17.24313827

**Authors:** Yanan Liu, Sara Jalali, Ridha Joober, Martin Lepage, Srividya Iyer, Jai Shah, David Benrimoh

## Abstract

Clinical course after first episode psychosis (FEP) is heterogeneous. Subgrouping longitudinal symptom trajectories after FEP would be useful for developing personalized treatment approaches, and being able to predict these trajectories at baseline would facilitate individual-level treatment planning. We utilized k-means clustering to identify distinct clusters of 411 FEP patients based on longitudinal positive and negative symptom patterns. Ridge logistic regression was then used to identify predictors of cluster membership using baseline data. Three clusters were identified, demonstrating unique demographic, clinical and treatment response profiles. Cluster 1 exhibits lower positive and negative symptoms (LS), lower antipsychotic dose, and relatively higher affective psychosis; Cluster 2 shows lower positive symptoms, persistent negative symptoms (LPPN), and intermediate antipsychotic doses; Cluster 3 presents persistently high levels of both positive and negative symptoms (PPNS), as well as higher antipsychotic doses. We effectively predicted patients’ cluster membership (AUC of 0.74). The most important predictive features included contrasting trends of apathy, affective flattening, and anhedonia for the LS and LPPN clusters. Global hallucination severity, positive thought disorder and manic hostility predicted PPNS. These results help parse the heterogeneity of FEP trajectories and may facilitate the development of personalized treatment approaches tailored to cluster characteristics.

## Introduction

Schizophrenia and other psychotic disorders affect roughly 1% of the population and impose significant disability and suffering on patients and caregivers ^1–4^. Major symptom categories leading to suffering and functional impairment include positive (e.g. hallucinations, delusions, disorganization) and negative (e.g. alogia, avolition, apathy) symptoms. Early intervention after the first episode of psychosis (FEP) has been identified as critical for improving outcomes for both negative and positive symptoms ^5^. However, the impact of early intervention services over regular treatment is still uncertain and parsing heterogeneity is necessary to improve treatment ^6^. Furthermore, the trajectories and treatment responsiveness of positive and negative symptoms show stark differences. Positive symptoms respond better to antipsychotic medication than negative symptoms, but may worsen over time and may be a factor leading to hospitalizations when insufficiently treated ^7^. Negative symptoms are less responsive to medication and generally more persistent ^8,9,10,11^; however, aspects of negative symptoms can improve after the first episode of psychosis and are responsive to psychological and psychosocial treatment ^12,13^. Overall, there is significant heterogeneity in symptomatology, illness course and neurobiological signatures in FEP^14^.

In order to improve personalized treatment planning and the development of novel treatments targeting specific symptom trajectories, it would be of value to determine if trajectories of positive and negative symptoms after FEP could be parsed into coherent subgroups. Furthermore, it would be useful to determine if membership in one of these putative subgroups could be predicted based on data available near the start of treatment. This would enable clinicians to proactively allocate resources and modify treatment plans in a manner that is patient- or person-centered. For example, if those at risk of treatment-resistant schizophrenia could be identified, more intensive interventions (such as intensive case management, psychosocial treatments, or early introduction of clozapine) could be considered in order to reduce the risk of the development of treatment resistance ^15^. In addition, the identification and characterization of symptom trajectory subgroups would provide important directions for future research aimed at developing subgroup-specific treatments or treatment protocols- a step towards personalized care.

While previous studies have looked at illness trajectory at a group level, or compared treatment-resistant schizophrenia with non-resistant groups, there is less research on symptom trajectory subgroups during treatment for a first episode psychosis ^16–18^. Here, we set out to determine if clear symptom trajectory subgroups exist after first episode psychosis, and if these can be predicted using baseline data. We employed a machine learning approach, K-means clustering, which has shown success in generating clusters in schizophrenia in previous work ^19^. We explored the differences in initial diagnoses, demographics, and onset history between the resulting symptom trajectory subgroups in order to better characterize them, and determined which presenting symptoms were most predictive of the subsequent trajectory subgroup membership. Furthermore, we explored how these subgroups interacted with treatment and treatment adherence in order to assess their relationship to treatment resistance.

## Method

### 1. Dataset and measures

The dataset for this study comes from the Prevention and Early Intervention for Psychosis Program (PEPP) in Montreal, Canada, which serves individuals experiencing FEP. The dataset includes 695 individuals aged 14 to 35 years (with an mean age of 23.64 ± 4.77 years); 69.9% are male. Data were collected from 2003-2018. Patients with an IQ below 70, organic brain damage, pervasive developmental disorders, or epilepsy were excluded. Eligible participants should not have had solely substance-induced psychosis and should have received antipsychotic treatment for no more than one month prior to being admitted to the program^20^.

This longitudinal dataset spans nine time points over two years, including patient evaluations and psychiatric treatments. Initially, at baseline, evaluators recorded patient demographics and assessed life history using the Course of Onset and Relapse Schedule (CORS) and Topography of Psychotic Episode (TOPE)^21^. Subsequently, at months 1, 2, 3, 6, 9, 12, 18, and 24, evaluators conducted semi-structured interviews to assess patient symptoms: positive and negative psychotic symptoms were evaluated using the Scale for the Assessment of Positive Symptoms (SAPS)^22^ and the Scale for the Assessment of Negative Symptoms (SANS)^20^. Furthermore, antipsychotic treatment dosage and patient adherence were recorded at each time point, with adherence^23^ levels of 0%, 25%, 50%, 75%, and 100% corresponding to never, very infrequent, sometimes, quite often, and always.

Antipsychotics were standardized into chlorpromazine equivalents(CE). The dosage of each antipsychotic was multiplied by a specific factor and the resulting converted doses were aggregated for each patient at each timepoint. The multiplication factor for each drug was established through previous studies ^24–35^ and is detailed in the supplementary materials. For injectable medications, we convert the dosage to an equivalent daily dose (total dose injected expressed as CE which was then divided by days between administrations) to ensure comparability with oral medications.

#### 1.1 Data Imputation

We employed the MissForest^36^ package for data imputation. Missing data was imputed for patients with at least 5 timepoints (i.e. > 50%) of negative and positive symptom scores (n = 411). We also conducted clustering on patients who had SAPS and SANS scores across all nine time points (n=122; complete case approach) as a sensitivity analysis (see supplementary materials).

### 2. Clustering Analysis

#### 2.1 Clustering approach and selection of cluster number

To classify patients into distinct clusters reflecting diverse positive and negative symptom trajectories, we applied the K-means^37^ clustering algorithm on trajectories of SAPS and SANS across all time points. This was our primary analysis. This unsupervised machine learning technique divides the data into k clusters according to their similarity in features. We tested k values ranging from 2 to 9, and determined the optimal number of clusters by considering the balance between inertia and silhouette score. Inertia represents the sum of squared distances of samples to their closest cluster center^38^; lower inertia signifies more compact clusters. Silhouette score^39^, on the other hand, quantifies the distance between intra-cluster and nearest inter-cluster distances; a higher silhouette score indicates more distinct and cohesive clusters. We calculated the ratio of inertia to silhouette score for each number of clusters and selected the number of clusters corresponding to the smallest ratio, striking a balance between cluster compactness and separation (see supplementary materials for further discussion).

#### 2.2 Cluster Membership Prediction

In order to explore the potential of the identified clusters for treatment planning in future studies, and to further characterize the clusters, in a secondary analysis we predicted patients’ cluster membership using supervised machine learning, with 40 features available at study entry, including demographics, onset history, baseline evaluations and presenting diagnoses (see Table 1). Features were selected from the dataset in order to cover demographics, onset history variables known to predict outcome, such as duration of untreated psychosis, and all the major symptom categories available at baseline. Baseline diagnostic categories were included because this information would be available after an initial assessment, the point in time where the prediction of this model would be used.

**Table 1.**
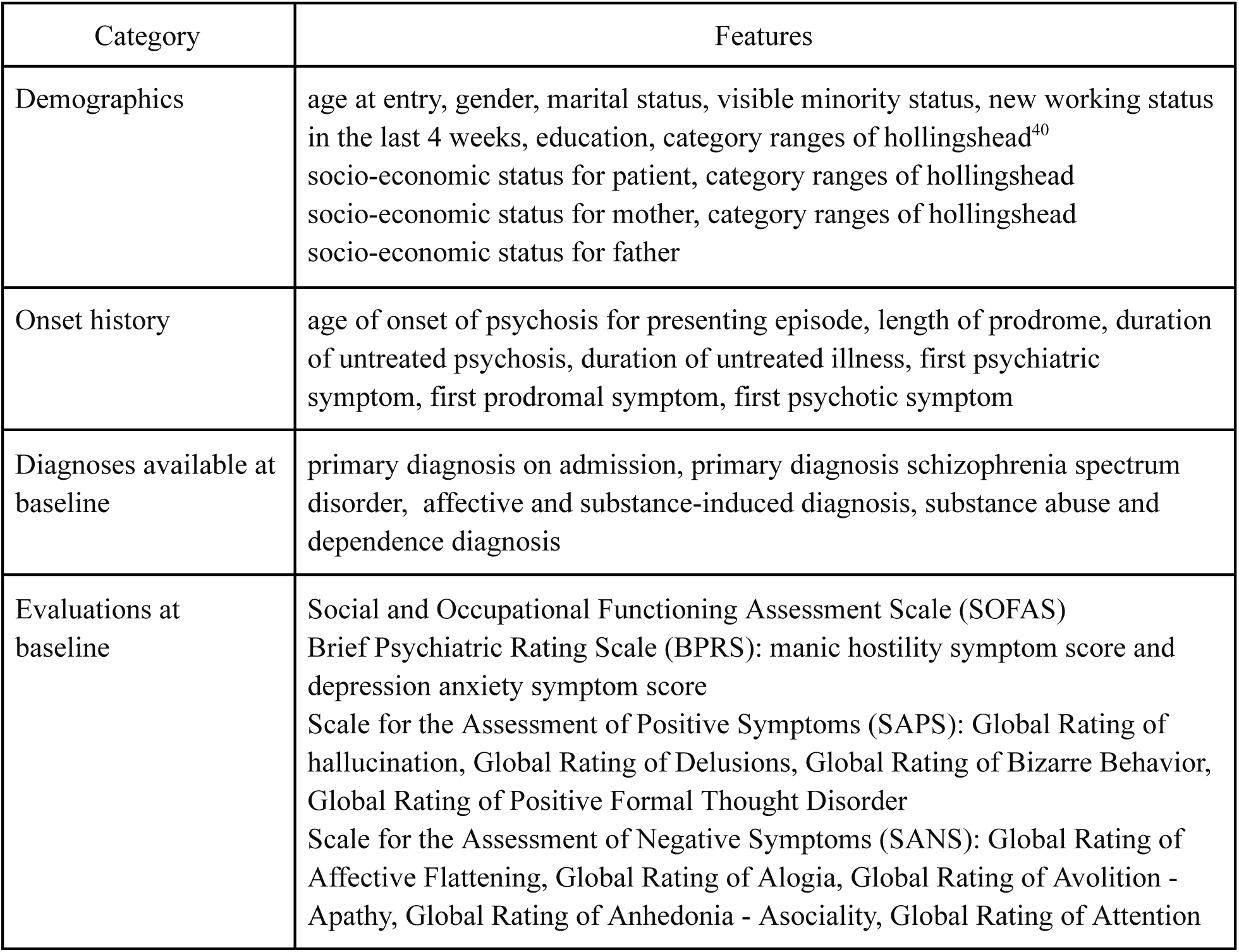
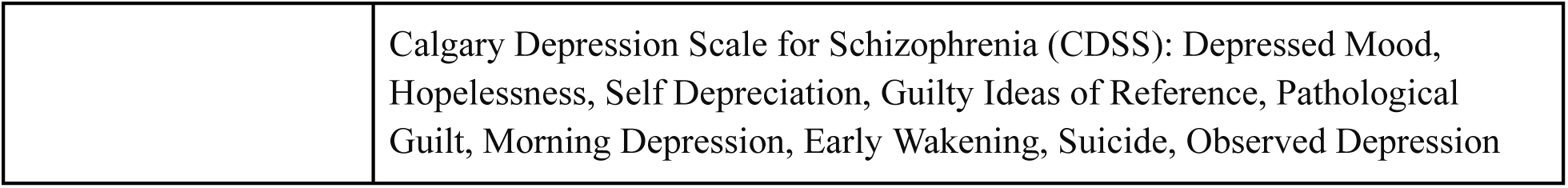
Baseline features for cluster membership prediction.

We applied one-hot encoding to transform categorical variables, and standard scaling to normalize numerical variables^38^. Following this preprocessing, we divided the datasets into training sets (80%) and testing sets (20%), stratified by cluster. We then utilized multiclass logistic regression^41^ with L2 penalty^42^ to predict cluster membership, and we used three-fold gridsearch to decide the optimal regularization strength, which helps prevent overfitting by penalizing large coefficients. The efficacy of the logistic regression model in predicting cluster membership was confirmed through a nonparametric permutation test. We randomly shuffled the cluster membership labels 1000 times, predicted the cluster membership of test sets for each permutation, and recorded the Accuracy (ACC) and Area Under the Receiver Operating Characteristic Curve (AUC). By comparing the AUC of the actual prediction to this distribution, we obtained a p-value to validate whether the prediction is successful. Additionally, we utilized the SHapley Additive exPlanations (SHAP) ^43^ package to characterize the feature importance in influencing the predicted cluster memberships.

### 3. Analysis of medication dosage and adherence over time

In an exploratory analysis, we characterized chlorpromazine equivalent trajectories over time across identified clusters to assess whether clinicians treated patients differently across these clusters. This may suggest an implicit recognition of differences in patient clinical presentations among clusters. In this observational dataset, we also preliminarily assessed the relationship of symptom severity with chlorpromazine equivalent over time. This also allowed us to examine if patients in different clusters were admitted at similar doses and, if dosing diverged over time, how this may have related to changes in symptoms. Finally, we examined medication adherence within each cluster to better understand treatment practices.

## Results

### 1. Overall sample characteristics

Table 2 lists the demographic data for all the participants in the analysis.

**Table 2.** Demographics for all patients. *Note: Cells including less than 5 values are censored to protect confidentiality*.

|  |  |
| --- | --- |
| Variables | N=411 |
| Age at entry, mean(std) | 23.35 (4.47) |
| Gender, N(%) |  |
| Male | 286 (69.6%) |
| Female | 125 (30.4%) |
| Marital status, N(%) |  |
| In a relationship | 368 (89.5%) |
| Not in a relationship | 39 (9.5%) |
| missing | 4 (1.0%) |
| Education level, N(%) |  |
| < high school | 138 (33.6%) |
| > high school | 257 (62.5%) |
| missing | 16 (3.9%) |
| SES<sup>1</sup>, N(%) |  |
| lower | 216 (52.5%) |
| middle | 71 (17.3%) |
| upper | 34 (8.3%) |
| missing | 90 (21.9%) |
| Ethnicity, N(%) |  |
| white | 260 (63.3%) |
| black | 38 (9.2%) |
| others | 99 (24.1%) |
| missing | 14 (3.4%) |

<sup>1</sup> Socioeconomic Status measured by the Hollingshead index<sup>40</sup>.

### 2. Three clusters identified as the optimal solution

As Fig. 1a demonstrates, the optimal number of symptom trajectory clusters is three, based on the ratio of inertia to silhouette score, which indicates both compactness and clear separation. We then calculated the size of each cluster (Fig. 1b). Cluster 1 comprises the majority with 228 patients, representing 55.47% of the total patient population. Clusters 2 and 3 contain 112 and 71 patients, respectively, accounting for 27.25% and 17.27% of the total. The trajectories these clusters represent is discussed below.

**Fig. 1.**
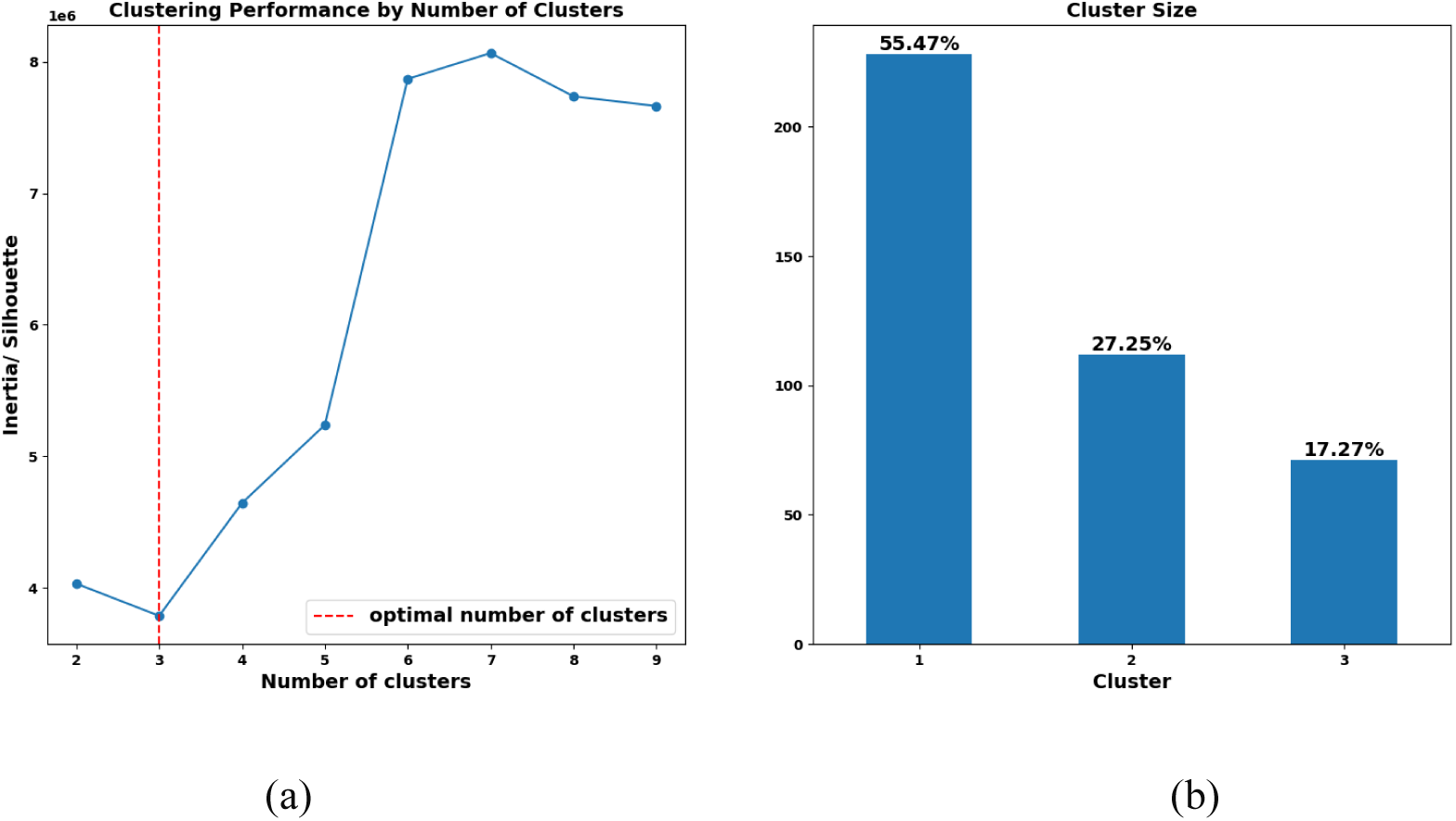
Optimal cluster number and the corresponding cluster size. (a) Clustering performance by number of clusters. X axis is the number of clusters, y axis is the ratio of inertia to silhouette score, which reflects the compactness within each cluster and the separation between clusters. The optimal number of clusters is 3, when the ratio of inertia to silhouette score reaches its minimum. (b) Cluster size. The number of patients in each cluster and their proportions. The trajectories these clusters represent are discussed below.

### 3. Distinct Symptom Trajectories in the Three Identified Clusters

The mean trajectory of SAPS and SANS total score for each group is illustrated in Fig. 2. We observed distinct patterns among the three clusters. In Cluster 1, both positive and negative symptoms start at a low level, decreasing even further, and stabilizing at a consistently low level. Consequently, we label cluster 1 "Low Symptoms" (LS). In contrast, cluster 2 displays a positive symptom pattern similar to cluster 1, while their negative symptoms start and persist at a higher level. Therefore, we label cluster 2 "Low Positive Persistent Negative" (LPPN). Cluster 3 demonstrates sustained higher levels of both positive and negative symptoms during the course of treatment, leading us to label this cluster as "Persistent Positive Negative Symptom" (PPNS). Importantly, while the three clusters have similar relative changes from baseline (Fig 2b), patients in the three clusters experience very different clinical outcomes by the end of their first two years of treatment in terms of absolute symptoms. As illustrated in Fig. 2b, positive symptom scores decreased notably and rapidly across all three groups, particularly within the initial two months of treatment in the PEPP program, whereas negative symptom scores demonstrated a slower decline over an extended period.

**Fig. 2.**
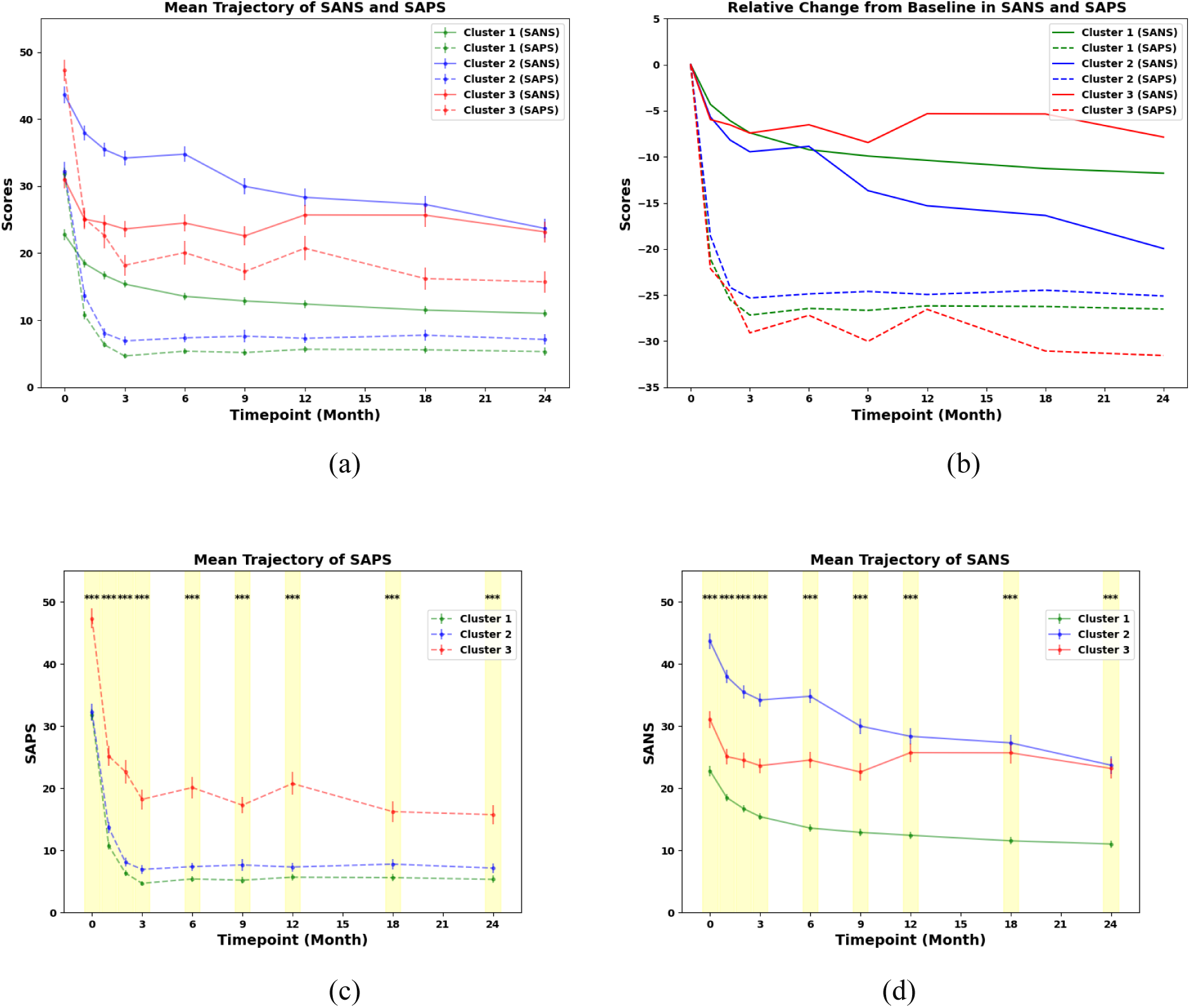
Symptom Score Trajectories. (a) Overall symptom trajectories with mean value and standard error. X axis is timepoint in months, y axis is the score of positive or negative symptom measured by SAPS or SANS. Solid line = negative symptoms; dashed line = positive symptoms. (b) Relative change from baseline. Each mean value was subtracted from the mean value at baseline to illustrate the relative change at each timepoint. (c) Positive symptom score trajectories with Kruskal-Wallis test significant levels, * means p < 0.05, ** means p <0.01, *** means p < 0.001 after bonferroni correction. (d) Negative symptom score trajectories, same as positive symptom. Color coding: green for cluster 1, blue for cluster 2, red for cluster 3.

As illustrated in Figures 2c and 2d, significant differences among the three clusters are evident for both positive and negative symptoms at each time point (see supplementary for supporting pairwise post-hoc analyses). It is important to note that there are significant differences in positive symptoms between cluster LS and PPNS as well as cluster LPPN and PPNS. For SANS, significant differences are observed between all pairs of clusters at each time point, except in the final year, where the difference between clusters PPNS and LPPN is no longer significant, suggesting higher levels of negative symptoms for both LPPN and PPNS once stabilized. (see supplementary material).

### 4. Cluster characterization

The demographics for each cluster is presented in Table 3.

**Table 3.** Demographics for clusters.

| Variables | Cluster 1 (N=228) | Cluster 2 (N=112) | Cluster 3 (N=71) |
| --- | --- | --- | --- |
| Age at entry, mean(std) | 24.06(4.53) | 22.47(4.33) | 22.44(4.08) |
| Gender, N(%) |  |  |  |
| Male | 145 (63.6%) | 92 (82.1%) | 49 (69.0%) |
| Female | 83(36.4%) | 20 (17.9%) | 22 (31.0%) |
| Marital status, N(%) |  |  |  |
| In a relationship | 199 (87.3%) | 106 (94.6%) | 63 (88.7%) |
| Not in a relationship | 27 (11.8%) | 5 (4.5%) | 7 (9.9%) |
| Missing | 2 (0.9%) | 1 (0.9%) | 1 (1.4%) |
| Education level, N(%) |  |  |  |
| < high school | 56 (24.6%) | 48 (42.8%) | 34 (47.9%) |
| > high school | 165 (72.4%) | 59 (52.7%) | 33 (46.5%) |
| missing | 7 (3.0%) | 5 (4.5%) | 4 (5.6%) |
| SES, N(%) |  |  |  |
| lower | 112 (49.1%) | 67 (59.8%) | 37 (52.1%) |
| middle | 42 (18.4%) | 17 (15.2%) | 12 (16.9%) |
| upper | 28 (12.3%) | <5 | <5 |
| missing | 46 (20.2%) | 24 (21.4%) | 20 (28.2%) |

| Ethnicity, N(%) |  |  |  |
| --- | --- | --- | --- |
| white | 146 (64.0%) | 67 (59.8%) | 47 (66.2%) |
| black | 16 (7.0%) | 13 (11.6%) | 9 (12.7%) |
| others | 58 (25.4%) | 27 (24.1%) | 14 (19.7%) |
| missing | 8 (3.5%) | 5 (4.5%) | 1 (1.4%) |

In addition to demographics, we analyzed the illness onset history for the three clusters as detailed in Table 4 and 5 (histograms in sFig. 4). We performed an ANOVA to test the age of onset, and applied the non-parametric Kruskal-Wallis test for length of prodrome, duration of untreated illness (DUI) and duration of untreated psychosis (DUP) due to their skewed distributions. For post-hoc analysis, t-tests were applied to age of onset, and Dunn’s tests were applied to the other three variables. We found significant differences in the age of onset, length of the prodrome and DUP among the three clusters. Post-hoc analysis revealed that, for age of onset and DUP, the differences were significant between cluster 1 and cluster 2, as well as between cluster 1 and cluster 3. For the length of the prodrome, the difference is significant between cluster 1 and cluster 2, with cluster 1 experiencing a shorter prodrome length.

**Table 4.** Cluster differences on age of onset (measure in years)

| Variable | Cluster 1 | Cluster 2 | Cluster 3 | ANOVA F | ANOVA p | C1 vs C2 | C2 vs C3 | C1 vs C3 |
| --- | --- | --- | --- | --- | --- | --- | --- | --- |
| ageonset | 23.31(4.71) | 21.54(4.28) | 21.31(3.87) | 8.94 | 0.000158*** | 0.000862** | 0.706 | 0.00121** |

*mean(std) for each cluster*
*C1 is cluster 1, C2 is cluster 2, C3 is cluster 3 for post-hoc analysis*
*\* means $p < 0.05$ , \*\* means $p < 0.01$ , \*\*\* means $p < 0.001$ with bonferroni correction*

**Table 5.** Cluster differences on duration of untreated illness (DUI), duration of untreated psychosis (DUP) and length of prodrome (PL) (measures in weeks)

| Variable | Cluster 1 | Cluster 2 | Cluster 3 | KW H | KW p | C1 vs C2 | C2 vs C3 | C1 vs C3 |
| --- | --- | --- | --- | --- | --- | --- | --- | --- |
| PL | 23.50(79.18) | 52.21(151.00) | 32.43(128.93) | 13.67 | 0.00107** | 0.000424** | 0.403 | 0.0395 |
| DUI | 243.43(352.57) | 258.71(421.43) | 243.57(362.86) | 1.82 | 0.402 | NA | NA | NA |
| DUP | 14.00(35.27) | 26.86(43.39) | 20.86(42.14) | 16.29 | 0.00029*** | 0.000382** | 0.87 | 0.0046* |

*median(IQR) for each cluster due to their skewness*
*C1 is cluster 1, C2 is cluster 2, C3 is cluster 3 for post-hoc analysis*
*\* means $p < 0.05$ , \*\* means $p < 0.01$ , \*\*\* means $p < 0.001$ with bonferroni correction*

We next examined baseline diagnoses within each cluster, in order to ensure that the clustering was not simply replicating diagnostic differences. We first examined differences between schizophrenia spectrum diagnoses (SSD) and affective psychoses (Fig. 3). The proportion of SSD exhibited a progressive increase from cluster 1 to cluster 2 to cluster 3, while conversely, the prevalence of affective psychosis showed a declining trend. Substance-induced psychosis was exclusive to cluster 1 (n = 1). We also examined the distribution of primary diagnoses at program start in each cluster (sFig. 3). It is important to note that the most common diagnoses in all three clusters were SSD. As such, these clusters do not simply recapitulate diagnostic differences at baseline; rather, they demonstrate variations of symptom trajectories even within those diagnosed with SSD at baseline.

**Fig. 3.**
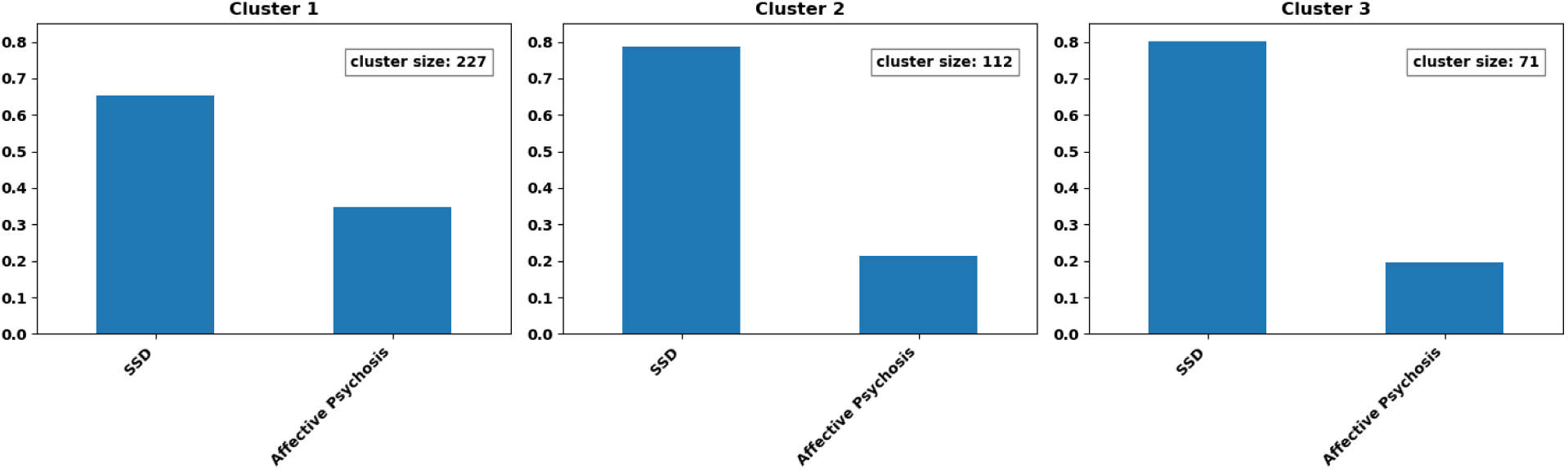
Diagnoses at baseline. Normalized bar plot of schizophrenia spectrum and affective psychosis for three clusters. (One patient in cluster 1 with substance-induced psychosis was excluded from this figure)

### 5. Prediction of cluster membership using data available at baseline

We successfully predicted patients’ cluster membership using baseline data. As shown in Fig. 4a, our model achieved an accuracy of 0.61, and AUC of 0.74 on the test set, and the permutation test indicates a p-value of 0, validating the prediction model. Subsequently, we identified the 15 most important features in the prediction of each cluster as depicted in Fig. 4b-d, which allows for further characterization of the clusters.

**Fig. 4.**
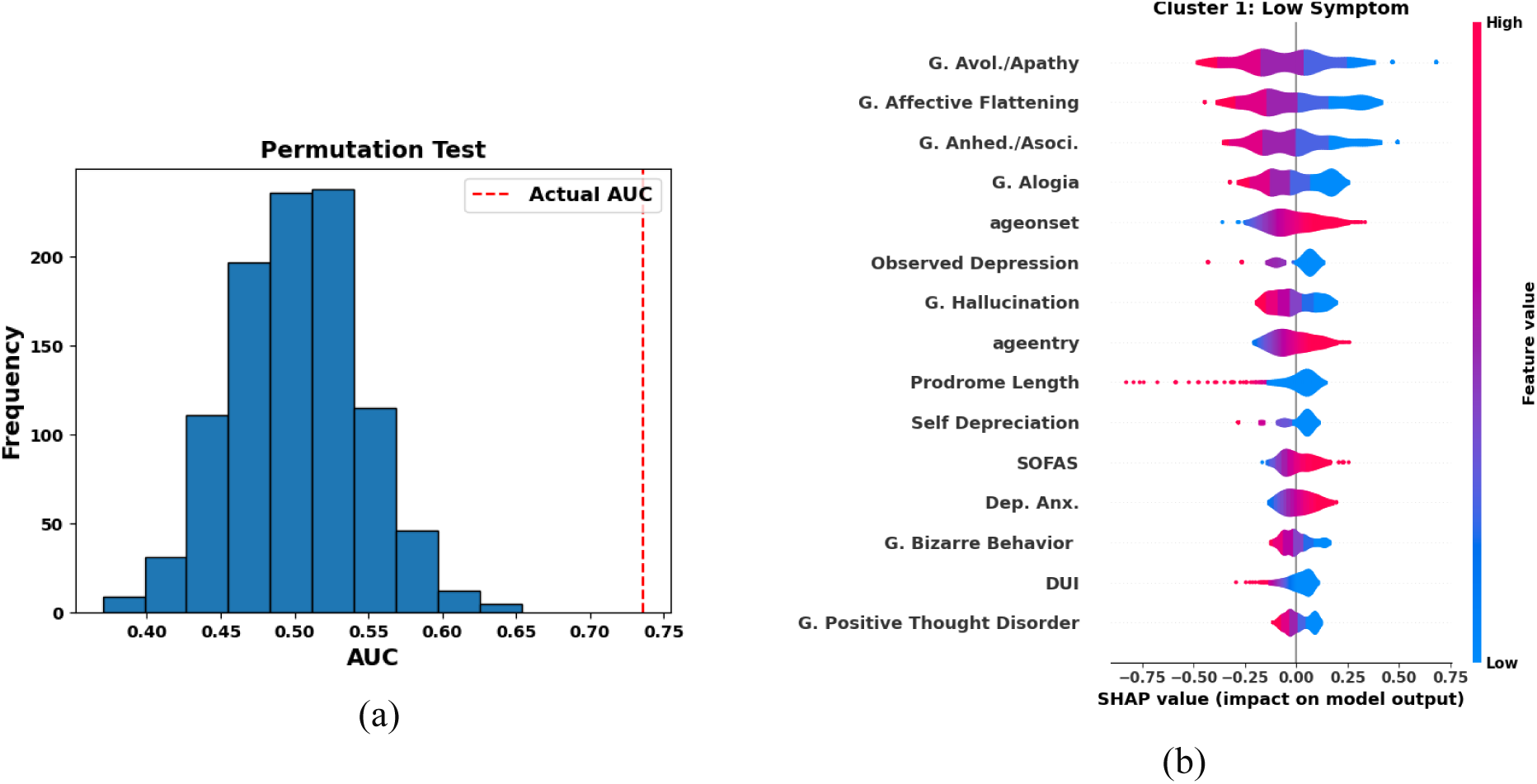
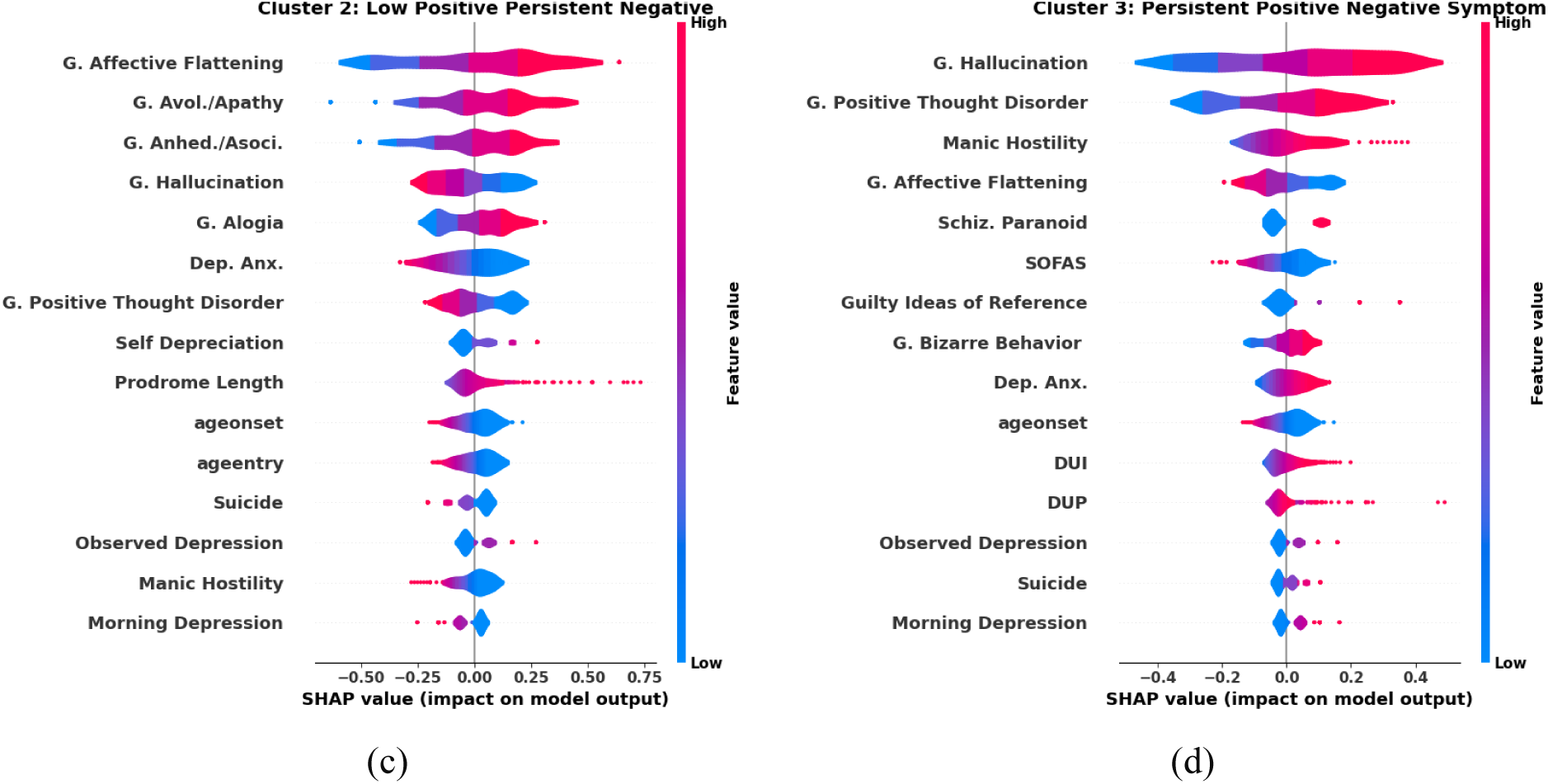
Cluster Membership Prediction and import features for predicting each cluster. (a) Permutation test. Histogram is the distribution of AUC for random prediction. Red vertical line is the actual prediction AUC. (b)-(d) The 15 most important features in the prediction for each cluster. X axis is SHAP value, representing the impact on model output of a specific feature, y axis is the feature name. Red indicates a higher value of the feature, and blue indicates a lower value.

For cluster LS, (Fig. 4b), the four most important features are global apathy, global affective flattening, global anhedonia, and global alogia; lower scores in these domains increase the likelihood of belonging to this cluster. In addition, both age of onset and age of entry are higher for cluster LS, while their length of prodrome and duration of untreated illness are shorter. Additionally, their functional level, measured by Social and Occupational Functioning Assessment Scale (SOFAS)^44^ at baseline, is higher. However, they exhibit higher depression and anxiety symptom scores, as measured by the BPRS-despite having lower levels of observed depression.

For cluster LPPN, as shown in Fig. 4c, the three most important features are the same as those observed in cluster LS. However, the trends differ; higher scores in these domains increase the likelihood of belonging to cluster LPPN. Patients in cluster LPPN tend to have a younger age of entry and onset, while they have a longer length of prodrome. In contrast to cluster LS, cluster LPPN shows lower depression and anxiety symptom scores-despite higher levels of observed depression, potentially consistent with the greater number of negative symptoms in this cluster which may appear depressive. Cluster LPPN tends to have lower positive thought disorder, which is an important distinction with cluster PPNS, suggesting biological or illness stage differences between the clusters. In addition, one of the most important predictive features for LPPN patients is a lower global hallucinations score, in contrast to PPNS.

For cluster PPNS as depicted in Fig. 4d, the most important distinguishing feature is global hallucination severity-PPNS patients have higher hallucinations, while the other two clusters show lower hallucinations. Global positive thought disorder and the manic hostility BPRS subscale are two key features as well; contrary to cluster LPPN, higher values indicate a stronger association with this cluster. Notably, cluster PPNS has a younger age of onset, a longer duration of untreated illness and untreated psychosis, implying delayed intervention despite relatively high symptom burden at intake. Besides, patients in cluster PPNS tend to have lower functional levels and a higher tendency toward suicidality. Consistent with sFig. 3, cluster PPNS is associated with higher paranoid schizophrenia. Importantly, despite baseline diagnosis being available to the model, it does not figure prominently in the prediction of trajectory cluster membership (aside from cluster 3), suggesting that individual patient symptoms, rather than overall diagnostic categories, are important for understanding patient progression in the identified clusters.

### 6. Differential Treatment Trajectories and Comparable Patient adherence Across Clusters

To further characterize the clusters and define their relationship with medication doses, we plotted chlorpromazine equivalents for each cluster. As shown in Figure 5a, all three clusters entered the service with equivalent antipsychotic doses, although differences emerged at the second month and increased over time. This is interesting, given that symptom scores changed substantially from the beginning of followup (Fig 2b), somewhat prior to when dose differences between clusters become the most marked (Fig 5a). Early treatment response appears similar between clusters in terms of relative changes (Fig 2b), but longer term absolute symptom levels are markedly different (Fig 2c, d), despite dose escalation in PPNS (Fig 5a).

**Fig. 5.**
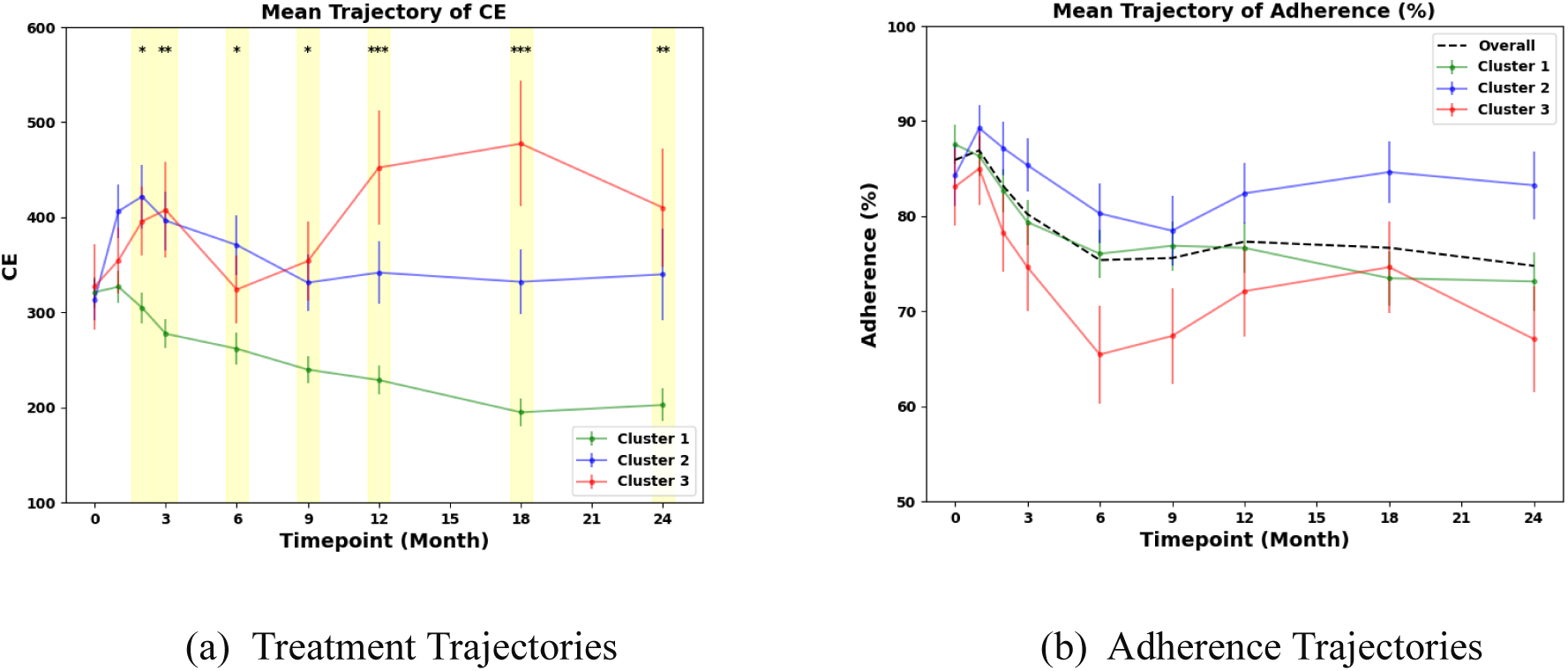
Treatment and adherence Trajectories. (a) Antipsychotic Dosage Trajectories. X axis is timpoint in months, y axis is the converted doses measured by chlorpromazine equivalents. Kruskal-Wallis test was conducted to evaluate the difference among three clusters.* notes significant levels, * means p < 0.05, ** means p <0.01, *** means p < 0.001 after bonferroni correction. (b) Mean adherence trajectories with standard error. Kruskal-Wallis test was also conducted at each timepoint, but none is significant. Adherence levels of 0%, 25%, 50%, 75%, and 100% means never, very infrequently, sometimes, quite often, and always taking medicines. Black dashed line is the mean adherence level for all patients.

For cluster LS, characterized by the mildest symptoms, the prescribed dose overall decreased to a lower level. For the LPPN, dosing increased and then decreased before stabilizing somewhat above the baseline level. For the PPNS, CE dose fluctuated in the first six months and then continued to increase for a year until at the last month, a decrease was noted (Fig 5a). Post-hoc analysis reveals that the primary difference is between the LS group and the others, with no significant difference between the LPPN and PPNS groups in terms of doses over time (sTable 3). Differences in clinician decisions regarding dosing in the three clusters provides some support for the clinical meaningfulness of our derived clusters.

We also plotted mean adherence levels trajectories over time (Fig. 5b). Overall, across all 3 clusters there was high adherence, suggesting that the distinct trajectories are not simply a result of varying treatment adherence. We calculated the proportion of patients who took 100% of their medications at each timepoint, per cluster. For cluster 1 this was 69.9%, for cluster 2 was 74.0%, for cluster 3 was 62.9%. For patients who always took at least 75% of their medication, the proportions for cluster 1 was 80.0%, for cluster 2 was 83.5%, and for cluster 3 was 72.2% (sTable 5, expanded across timepoints in sTable 6). No significant differences were observed among the three clusters at any time points (Fig. 5b, sTable 4), suggesting a negligible effect of adherence. However, it is noteworthy that cluster PPNS exhibits the lowest absolute adherence most of the time.

## Discussion

Using machine learning, we identified three clusters of psychotic symptom trajectories after a FEP, each characterized by unique demographics, illness histories, longitudinal symptom patterns and prescribed antipsychotic doses. Furthermore, we have demonstrated that it is possible to predict membership in a longitudinal symptom cluster using data available upon entry into a clinic. These findings may eventually allow clinicians to identify patients at risk for less favorable illness trajectories earlier in treatment, allowing for the modification of treatment plans and the assignment of clinical resources to better meet expected individual patient needs. In addition, the subgroups detailed here could serve as targets for the development of novel, personalized treatment approaches, based on further neurobiological and psychosocial research into the etiology of each subgroup. To our knowledge, ours is the first study to identify symptom trajectory clusters in a longitudinal first episode sample based on both positive and negative symptoms, and to predict membership in these clusters using symptom data available solely at baseline.

In this optic, it is interesting to examine findings from each cluster, and from differences between clusters, which may support improvement in treatment approaches or provide targets for further work. The first cluster, LS, characterized by low positive and negative symptoms and reduced dose of antipsychotic treatment, has a lower duration of untreated psychosis (DUP) - an important predictor of outcome ^5^.The third cluster, PPNS, displays the highest level of positive symptoms (concordant with them receiving higher antipsychotic doses than the LS cluster) and negative symptoms comparable to those in the LPPN group. Despite extensive treatment and reduction in symptoms, patients in PPNS continue to show persistently higher symptoms over time. This suggests a ceiling effect of treatment in this subgroup, and further suggests potential biological differences underlying positive symptoms between this cluster and the other two clusters which could be a fruitful target for further research.

Crucially, we were able to predict cluster membership using data at baseline, which raises the exciting possibility of secondary prevention approaches aimed at reducing the incidence of treatment resistance. This means that treatments developed for specific subgroups would not need to necessarily be targeted at that subgroup at the time when their symptoms are at their peak; rather, they could be developed with the aim of *preventing* progression along the predicted trajectory by intervention earlier in the illness course (and, potentially, prior to illness onset). This would be an approach similar to the one currently gaining traction in the treatment of Alzheimer’s dementia, where treatments seem most effective when targeting pathophysiological processes prior to when they generate maximal symptoms^45^.

In addition it is interesting to discuss the features most predictive of cluster membership. Clusters 1 and 2 had the same top three features- all dimensions of the negative symptoms, with different directions for each (less symptom intensity in cluster 1, more in cluster 2); this is consistent with the fact that between clusters 1 and 2, the main difference is the presence of negative symptoms. However, in cluster 3, the most important features are hallucinations and positive formal thought disorder. Poor function at baseline also predicts this cluster, and there is less affective flattening at baseline compared to the LPPN cluster (suggesting a different pattern of negative symptoms than LPPN, likely related to the more significant presentation of psychotic symptoms in this cluster). In addition, this is the only cluster with increased manic hostility (despite having lower incidence of affective psychosis, suggesting this score was tapping into a more cross-diagnostic measure of hostility) and bizarre behavior as key predictive symptoms. This suggests not simply more positive symptoms in this group- as would be expected given the persistence of positive symptoms is characteristic of cluster 3-but a more disorganized (in terms of thought and behavior) and less functional presentation. This phenotype being prognostic of poorer outcomes is consistent with some literature^46^ and merits further investigation. The importance of hallucinations as a defining feature of this poorer outcome cluster is especially interesting for future study. In recent work^47^ we have shown that delusions are more common and generally more stable in early psychosis and psychosis risk than hallucinations, meaning delusion presence might be less likely to differentiate between patients. In addition, hallucinations also often respond to treatment more rapidly than delusions^48^. As such, high hallucination severity even after initial treatment may represent a more severe psychosis phenotype and the mechanisms underlying this warrant further study. Finally, the increased suicidality scores predicting PPNS cluster membership at baseline is an important area for further exploration.

To be useful in potential treatment development, it would be important to identify neurobiological differences between our identified trajectory clusters. While this is not possible with our current data, the three clusters identified in this study align with the three biotypes identified by the B-SNIP Consortium^49^, who clustered patients based on neuropsychological measures and electrophysiological signals based on six laboratory tasks and validated their clustering in a follow-up study^19^. Their biotype 3, characterized by more affective psychosis, lower positive and negative symptom scores, and higher functional levels, is analogous to cluster LS in this study. Conversely, biotype 1, which has the opposite trends, aligns with cluster PPNS. They characterized the neuroanatomical deviation of their three biotypes with structural magnetic resonance imaging (sMRI) and found that the gray matter reduction in biotype 1 is the most widespread among the three clusters. Combined with previous studies which have identified two^50–52^ or three^53–55^ clusters of schizophrenia using neuroimaging, these findings support the presence of distinct biological mechanisms underlying different psychosis subtype clusters, and increases the probability of identifying meaningful neurobiological differences between them.

One of the strengths of this study was the use of a large dataset which captures data in the course of naturalistic care. This makes it more likely that the clusters identified here are representative of real clinical practice, and potentially generalizable to other clinical samples - and indeed, replication of these findings in other samples should be a focus of future work. One corresponding limitation to this study is that its observational nature means that individual differences in clinician prescribing or fluctuations in the availability of psychosocial treatments may have influenced symptom trajectories for some patients.

Following replication, future research might include prospective studies in which cluster membership prediction using the readily available clinical data used here is harnessed to predict response to treatment and guide treatment course (for example, by providing patients with poorer predicted symptom courses access to more intensive psychosocial support or more rapid consideration of clozapine). In addition, further data types, such as brain scans, computational parameters derived from tasks^56^ and genetic testing could be investigated as potential features for improving cluster membership prediction. Future work investigating underlying neurobiological and psychosocial differences between the LS and other clusters and between the LPPN and PPNS clusters could provide novel insights into the processes driving symptom presentation and treatment response. Finally, future work using computational phenotyping could be used to further identify latent states driving these differences^56^.

## Data Availability

All data produced in the present study are available upon reasonable request to the authors

