## supplementary material for "Subtyping First-Episode Psychosis based on Longitudinal Symptom Trajectories Using Machine Learning"

#### Results

##### 1. Optimal number of cluster

To determine the optimal number of clusters, we combined two criteria: inertia and silhouette score. Inertia measures within-cluster distance[1], with lower values indicating tighter, more compact clusters. As sFig. 1 shows, as the number of clusters increases, inertia decreases; however, an excessively high number of clusters can lead to overfitting, where each data point could end up in its own cluster in the most extreme case, rendering the clustering results meaningless.

The silhouette score evaluates how well-separated the clusters are by comparing the average distance between points within the same cluster to the average distance to points in the nearest cluster[1]. Higher silhouette scores reflect more distinct and well-separated clusters. As shown in sFig. 1, the silhouette scores for two and three clusters are similar, but inertia decreases sharply with more clusters. Considering both measures, we determined that the optimal number of clusters is three.

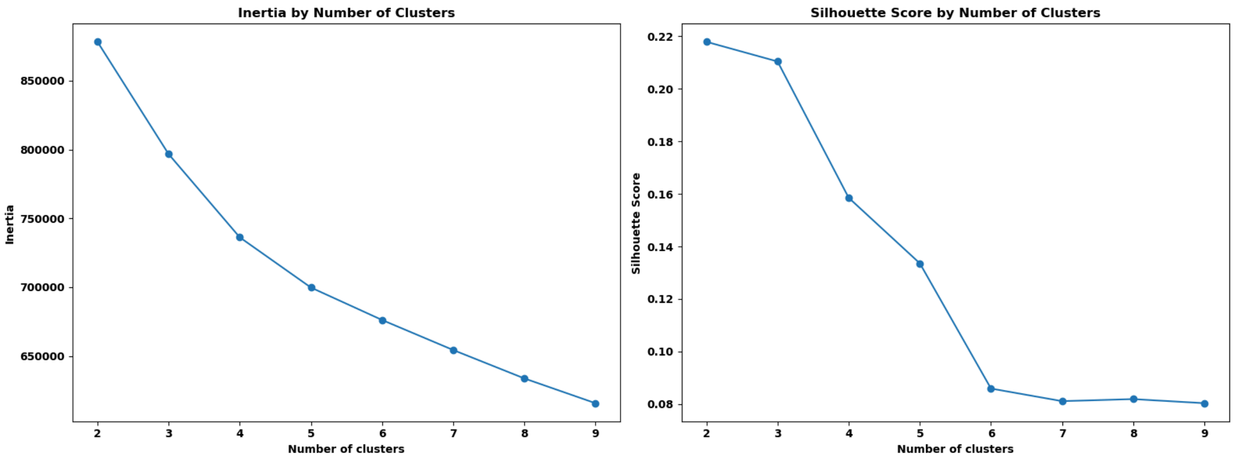


sFig. 1. Clustering number selection.

##### 2. Complete case clustering analysis

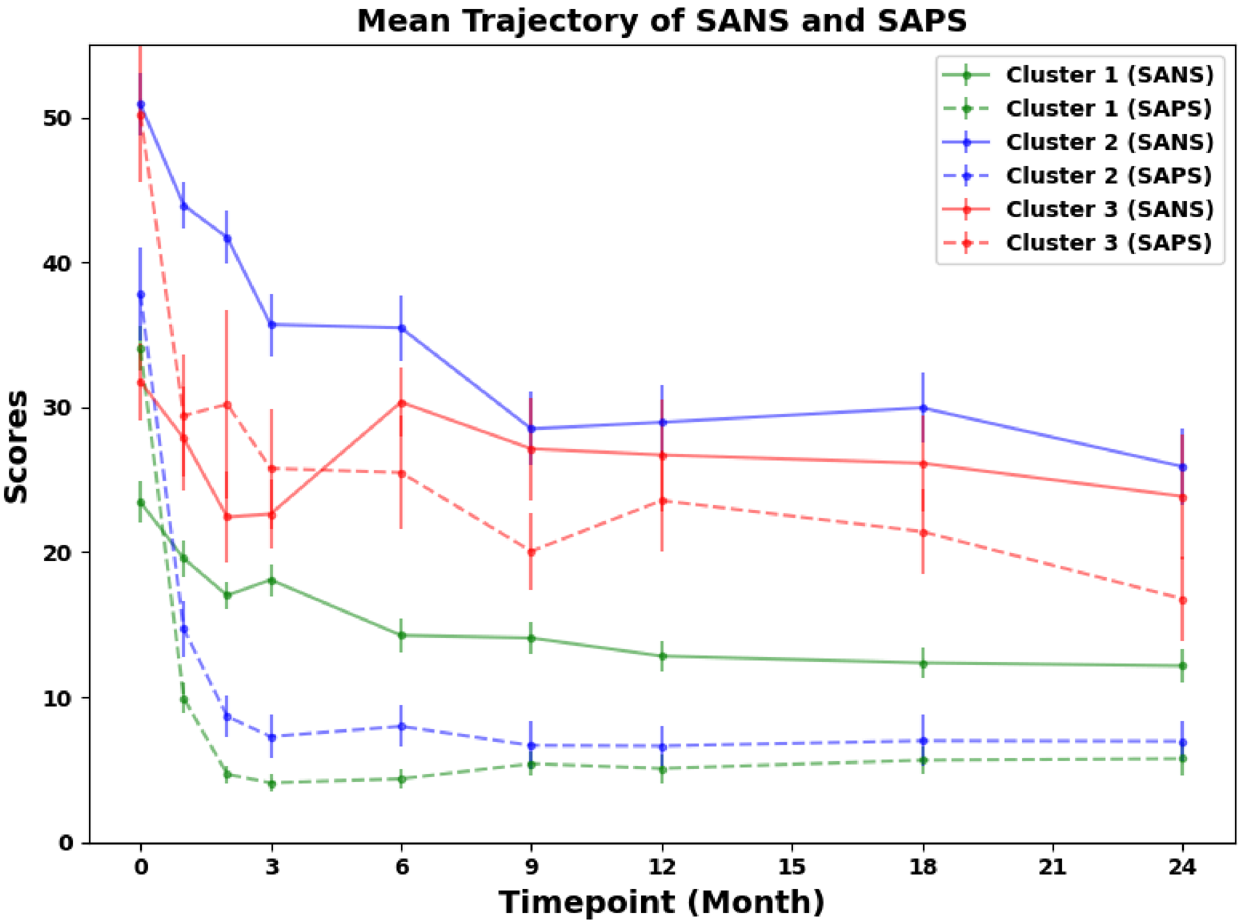


sFig. 2. Clustering results on un-imputed raw data. Patients with missing SAPS or SANS at any timepoint were excluded, resulting in a final sample of 122 patients. K-means clustering was applied to the remaining symptom trajectories to identify three clusters. The x-axis represents timepoints in months, and the y-axis indicates SAPS or SANS scores.

#### 3. Primary diagnosis distribution

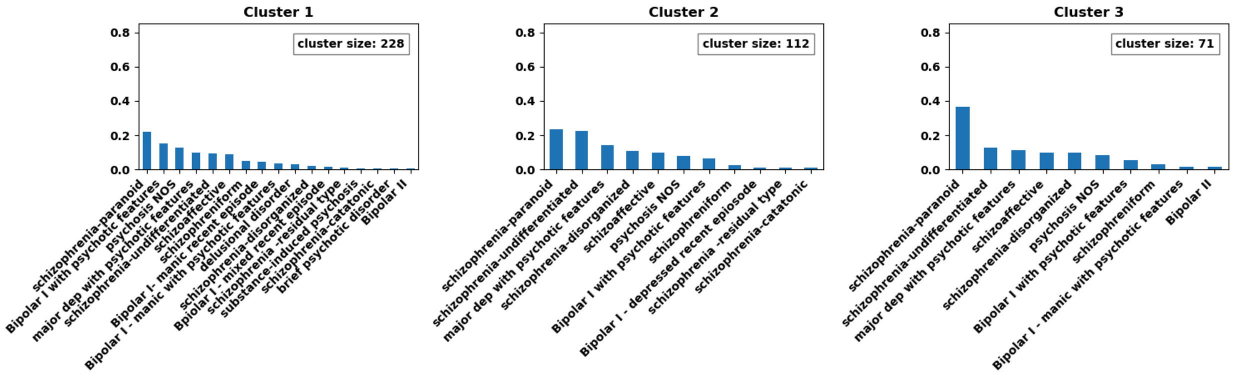


sFig3. Diagnoses at baseline. Normalized bar plot of primary diagnosis on admission for the three clusters.

#### 4. Distribution of prodrome length, DUI and DUP

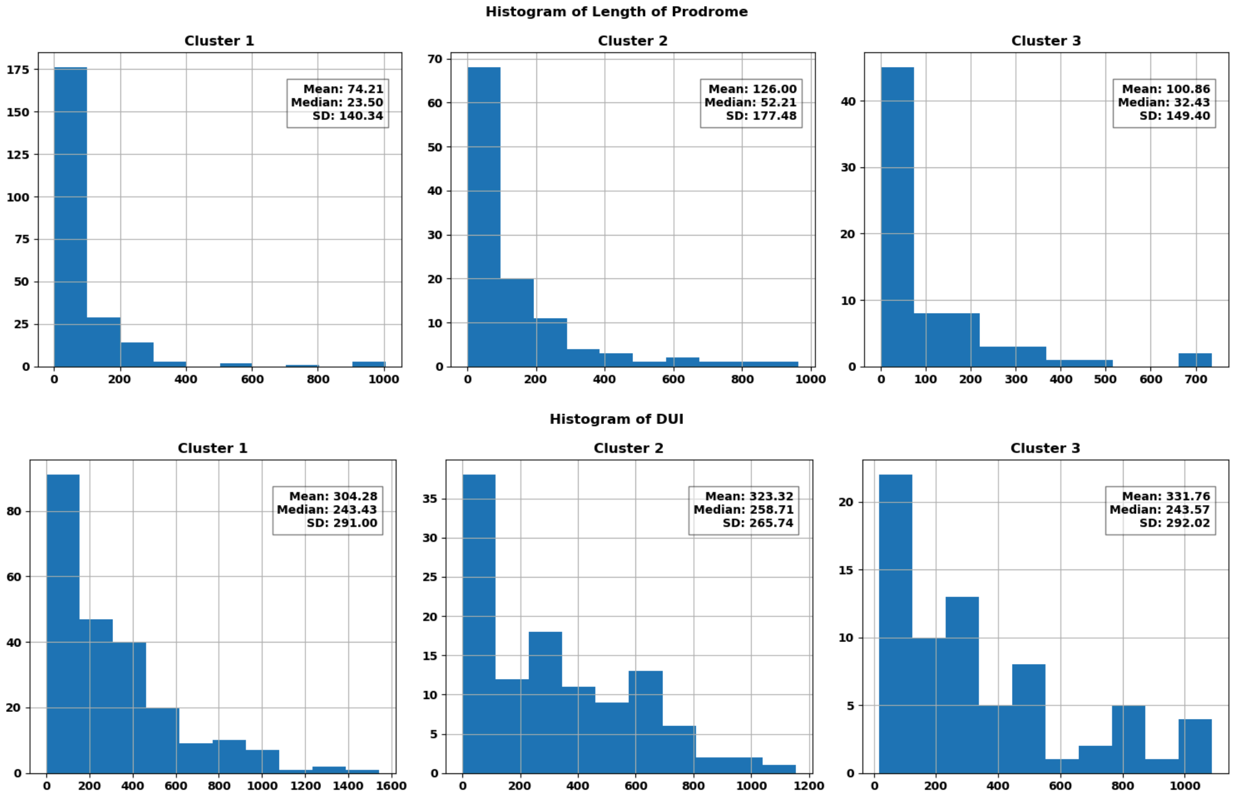

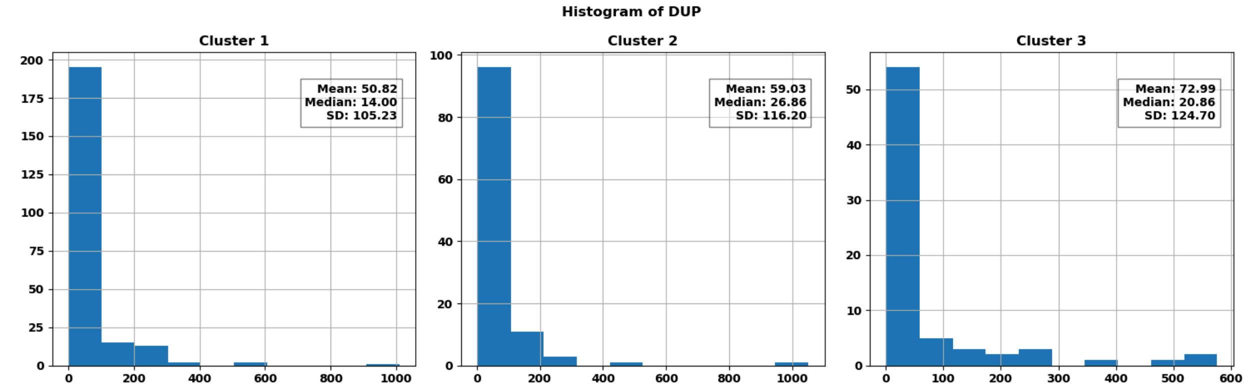


sFig4. Histogram of length of prodrome, DUI and DUP

#### 5. Statistical analysis for SAPS, SANS, CE and adherence

sTable 1. SAPS statistical analysis

| month | C1 mean(std, median, IQR) | C2 mean(std, median, IQR) | C3 mean(std, median, IQR) | KW H | KW p | C1-C2 | C2-C3 | C1-C3 |
|---|---|---|---|---|---|---|---|---|
| 0 | 31.83(13.73, 31.00, 18.00) | 32.23(14.76, 31.00, 19.00) | 47.28(13.45, 46.00, 16.50) | 59.84 | 1.01e-13*** | 0.823 | 6.3e-11*** | 7.08e-14*** |
| 1 | 10.74(9.04, 9.50, 14.00) | 13.65(9.24, 13.00, 13.00) | 25.15(13.68, 22.00, 15.50) | 72.27 | 2.02e-16*** | 0.00684 | 2.81e-08*** | 1.99e-17*** |
| 2 | 6.32(6.66, 4.00, 8.00) | 8.06(7.38, 6.00, 10.00) | 22.62(16.34, 19.00, 21.00) | 90.13 | 2.68e-20*** | 0.0277 | 9.06e-12*** | 2.49e-21*** |
| 3 | 4.66(5.71, 2.00, 8.00) | 6.91(7.16, 5.00, 10.00) | 18.18(13.31, 16.00, 14.50) | 83.85 | 6.18e-19*** | 0.00357 | 2.22e-09*** | 5.71e-20*** |
| 6 | 5.38(7.34, 2.00, 8.00) | 7.36(7.16, 5.50, 9.00) | 20.08(14.70, 18.00, 20.50) | 85.28 | 3.04e-19*** | 0.00118* | 7.34e-09*** | 3.31e-20*** |
| 9 | 5.18(7.35, 2.50, 8.00) | 7.62(9.54, 5.00, 10.25) | 17.25(10.74, 16.00, 13.00) | 90.69 | 2.03e-20*** | 0.00625 | 1.11e-10*** | 1.69e-21*** |
| 12 | 5.66(7.95, 3.00, 8.00) | 7.29(7.35, 5.50, 7.25) | 20.73(15.40, 18.00, 19.00) | 87.17 | 1.18e-19*** | 0.0058 | 3.74e-10*** | 1.01e-20*** |
| 18 | 5.59(8.74, 2.00, 7.00) | 7.76(9.05, 5.00, 7.25) | 16.21(14.14, 14.00, 15.00) | 59.13 | 1.45e-13*** | 0.0034 | 3.81e-06*** | 2.08e-14*** |
| 24 | 5.31(8.91, 2.00, 6.00) | 7.12(8.29, 4.00, 8.25) | 15.72(13.27, 12.00, 17.50) | 56.99 | 4.22e-13*** | 0.00175* | 1.62e-05*** | 7.98e-14*** |

*C1 is cluster 1, C2 is cluster 2, C3 is cluster 3*
*KW is Kruskal–Wallis test*
** means p < 0.05 , ** means p < 0.01, *** means p < 0.001 with bonferroni correction*

sTable 2. SANS statistical analysis

| month | C1 mean(std, median, IQR) | C2 mean(std, median, IQR) | C3 mean(std, median, IQR) | KW H | KW p | C1-C2 | C2-C3 | C1-C3 |
|---|---|---|---|---|---|---|---|---|
| 0 | 22.79(12.46, 21.00, 16.00) | 43.64(13.23, 44.00, 19.25) | 31.03(11.84, 29.00, 15.00) | 135.65 | 3.5e-30*** | 4.48e-31*** | 7.47e-07*** | 1.57e-05*** |
| 1 | 18.48(9.09, 18.00, 11.00) | 37.96(11.79, 37.00, 16.25) | 25.07(10.61, 24.00, 13.00) | 163.20 | 3.64e-36*** | 2.88e-37*** | 3.44e-09*** | 2.29e-05*** |
| 2 | 16.70(8.71, 16.00, 13.00) | 35.46(11.18, 35.50, 13.25) | 24.49(10.29, 26.00, 13.50) | 162.20 | 6.02e-36*** | 1.31e-36*** | 3.61e-07*** | 4.42e-07*** |
| 3 | 15.39(8.58, 15.00, 13.00) | 34.19(11.35, 33.00, 15.50) | 23.61(10.08, 25.00, 11.00) | 166.75 | 6.18e-37*** | 2.91e-37*** | 1.8e-06*** | 3.81e-08*** |
| 6 | 13.56(8.77, 12.00, 12.00) | 34.78(11.83, 32.00, 16.00) | 24.51(10.87, 24.00, 16.00) | 190.56 | 4.18e-42*** | 1.93e-41*** | 1.44e-05*** | 3.92e-11*** |
| 9 | 12.87(9.03, 12.00, 11.25) | 29.97(13.29, 29.00, 20.00) | 22.58(12.07, 20.00, 13.50) | 133.56 | 9.93e-30*** | 4.98e-29*** | 0.00141* | 3.03e-09*** |
| 12 | 12.40(9.03, 11.00, 11.00) | 28.33(14.12, 27.00, 19.25) | 25.70(12.62, 25.00, 14.50) | 135.56 | 3.65e-30*** | 3.25e-25*** | 0.456 | 1.51e-15*** |
| 18 | 11.51(9.14, 10.00, 11.00) | 27.28(13.73, 25.00, 19.00) | 25.68(15.17, 25.00, 20.50) | 131.67 | 2.55e-29*** | 4.14e-25*** | 0.308 | 2.01e-14*** |
| 24 | 11.00(9.00, 9.00, 10.00) | 23.69(15.17, 21.00, 20.00) | 23.17(13.21, 21.00, 16.00) | 93.60 | 4.74e-21*** | 2.75e-16*** | 0.792 | 4.4e-13*** |

sTable 3. CE statistical analysis

| month | C1 mean(std, median, IQR) | C2 mean(std, median, IQR) | C3 mean(std, median, IQR) | KW H | KW p | C1-C2 | C2-C3 | C1-C3 |
|---|---|---|---|---|---|---|---|---|
| 0 | 321.11(242.20, 275.07, 314.59) | 313.41(228.66, 265.88, 374.95) | 327.05(380.12, 250.00, 257.60) | 0.63 | 0.731 | 0.954 | 0.516 | 0.439 |
| 1 | 326.80(248.85, 254.50, 337.76) | 406.26(303.32, 348.95, 276.79) | 354.58(293.55, 300.00, 374.95) | 5.07 | 0.0792 | 0.0246 | 0.204 | 0.625 |
| 2 | 304.80(246.51, 250.00, 317.71) | 421.81(356.18, 333.65, 256.67) | 395.71(305.42, 375.00, 409.48) | 11.88 | 0.00263* | 0.00186 | 0.756 | 0.0218 |
| 3 | 277.41(234.98, 250.00, 322.88) | 396.35(326.02, 333.31, 256.67) | 407.62(423.26, 312.50, 384.17) | 16.00 | 0.000336** | 0.000217** | 0.564 | 0.0126 |
| 6 | 261.67(253.50, 223.21, 274.99) | 370.74(331.71, 333.30, 317.66) | 323.99(299.31, 250.00, 413.67) | 12.53 | 0.00191* | 0.000473* | 0.191 | 0.132 |
| 9 | 239.67(209.14, 223.21, 249.84) | 331.16(312.66, 312.50, 273.77) | 353.81(353.44, 250.00, 333.30) | 10.57 | 0.00506* | 0.00584 | 0.939 | 0.0153 |
| 12 | 228.71(224.71, 187.50, 239.18) | 341.77(349.92, 250.00, 316.60) | 452.22(506.27, 333.30, 318.56) | 22.10 | 1.59e-05*** | 0.00324 | 0.0988 | 1.41e-05*** |
| 18 | 194.77(218.98, 161.45, 291.75) | 332.04(361.55, 250.00, 321.55) | 477.37(554.50, 334.82, 541.85) | 30.65 | 2.21e-07*** | 0.000114** | 0.148 | 1.01e-06*** |
| 24 | 202.49(260.86, 125.00, 270.76) | 339.90(513.09, 223.21, 437.50) | 410.05(520.28, 250.00, 507.26) | 16.23 | 0.000299** | 0.00812 | 0.207 | 0.000256** |

sTable 4. Adherence statistical analysis (not-applicable patients were excluded)

| month | C1 mean(std, median, IQR) | C2 mean(std, median, IQR) | C3 mean(std, median, IQR) | KW H | KW p | C1-C2 | C2-C3 | C1-C3 |
|---|---|---|---|---|---|---|---|---|
| 0 | 87.56(30.81, 100.00, 0.00) | 84.26(33.77, 100.00, 0.00) | 83.09(33.30, 100.00, 6.25) | 1.96 | 0.376 | 0.539 | 0.444 | 0.169 |
| 1 | 86.32(30.43, 100.00, 0.00) | 89.25(25.49, 100.00, 0.00) | 85.00(30.87, 100.00, 25.00) | 0.81 | 0.667 | 0.692 | 0.369 | 0.506 |
| 2 | 82.70(33.89, 100.00, 6.25) | 87.16(29.55, 100.00, 0.00) | 78.26(34.28, 100.00, 25.00) | 5.68 | 0.0585 | 0.221 | 0.0172 | 0.105 |
| 3 | 79.37(35.52, 100.00, 25.00) | 85.36(29.48, 100.00, 25.00) | 74.64(37.99, 100.00, 50.00) | 3.62 | 0.164 | 0.213 | 0.0614 | 0.302 |
| 6 | 76.04(37.22, 100.00, 25.00) | 80.28(32.82, 100.00, 25.00) | 65.44(42.33, 100.00, 75.00) | 4.75 | 0.0931 | 0.48 | 0.0327 | 0.0757 |
| 9 | 76.88(37.73, 100.00, 25.00) | 78.47(36.92, 100.00, 25.00) | 67.39(41.64, 100.00, 75.00) | 3.77 | 0.152 | 0.804 | 0.0767 | 0.0753 |
| 12 | 76.66(38.31, 100.00, 25.00) | 82.40(31.75, 100.00, 25.00) | 72.10(39.88, 100.00, 50.00) | 1.82 | 0.404 | 0.447 | 0.178 | 0.392 |
| 18 | 73.47(40.16, 100.00, 50.00) | 84.64(31.91, 100.00, 0.00) | 74.63(39.77, 100.00, 50.00) | 5.35 | 0.0689 | 0.0227 | 0.124 | 0.784 |
| 24 | 73.13(40.77, 100.00, 50.00) | 83.24(33.96, 100.00, 12.50) | 67.06(43.95, 100.00, 100.00) | 5.94 | 0.0512 | 0.0631 | 0.0207 | 0.346 |

sTable 5. Adherence distribution by subtype (not-applicable patients were excluded)

| Adherence (%) | 0% (never) | 25% (very infrequent) | 50% (sometimes) | 75% (quite often) | 100% (always) |
|---|---|---|---|---|---|
| **subtype 1** | 14.6% | 2.3% | 4.2% | 9.1% | 69.9% |
| **subtype 2** | 9.5% | 2.6% | 4.4% | 9.4% | 74.0% |
| **subtype 3** | 17.0% | 4.4% | 6.4% | 9.2% | 62.9% |
| **Total** | 13.6% | 2.7% | 4.6% | 9.2% | 69.8% |

sTable 6. Adherence distribution by timepoints

| Adherence timepoint | 0% (never) | 25% (very infrequent) | 50% (sometimes) | 75% (quite often) | 100% (always) | Not Applicable |
|---|---|---|---|---|---|---|
| 0 | 40 (9.7%) | 9 (2.2%) | 12 (2.9%) | 15 (3.6%) | 325 (79.1%) | 10 (2.4%) |
| 1 | 30 (7.3%) | 7 (1.7%) | 12 (2.9%) | 36 (8.8%) | 299 (72.7%) | 27 (6.6%) |
| 2 | 40 (9.7%) | 14 (3.4%) | 22 (5.4%) | 26 (6.3%) | 302 (73.5%) | 7 (1.7%) |
| 3 | 51 (12.4%) | 8 (1.9%) | 20 (4.9%) | 51 (12.4%) | 273 (66.4%) | 8 (1.9%) |
| 6 | 61 (14.8%) | 13 (3.2%) | 27 (6.6%) | 50 (12.2%) | 242 (58.9%) | 18 (4.4%) |
| 9 | 65 (15.8%) | 12 (2.9%) | 19 (4.6%) | 40 (9.7%) | 247 (60.1%) | 28 (6.8%) |
| 12 | 58 (14.1%) | 11 (2.7%) | 18 (4.4%) | 42 (10.2%) | 249 (60.6%) | 33 (8.0%) |
| 18 | 59 (14.4%) | 13 (3.2%) | 15 (3.6%) | 30 (7.3%) | 242 (58.9%) | 52 (12.7%) |
| 24 | 64 (15.6%) | 7 (1.7%) | 14 (3.4%) | 26 (6.3%) | 217 (52.8%) | 83 (20.2%) |

#### 6. Linear mixed model of treatment and adherence

To assess the effect of treatment and patient adherence on symptoms while considering individual differences, we applied a linear mixed effects model. The regression model examines the relationship between the outcome variable which could be SAPS or SANS scores and several key predictors: chlorpromazine equivalent (CE), adherence, their interaction and timepoint. Additionally, the model includes a random intercept for each participant to account for individual differences that may affect the baseline scores. sTable 7 presents an outline of the fixed effects considered in the analysis. Time emerges as the primary factor, exerting a highly significant influence on both SAPS and SANS. In line with our expectations, treatment effect (CE) and adherence show a stronger impact on SAPS than on SANS. Importantly, the effect of CE was stronger than that of adherence, and there was no interaction between CE and adherence, suggesting that dosing does not systematically affect adherence rates.

sTable 7. Fixed effect of linear mixed model

| | SAPS | | SANS | |
|---|---|---|---|---|
| fixed effect | F value | p value | F value | p value |
| CE | 18.83 | 1.47e-05*** | 7.62 | 5.82e-03** |
| Adherence | 5.72 | 6.72e-04*** | 2.72 | 4.29e-02* |
| CE * Adherence | 0.31 | 8.16e-01 | 1.26 | 2.86e-01 |
| timepoint | 311.12 | 5.10e-66*** | 267.38 | 2.66e-57*** |

### Discussion

It is also interesting to examine clinician prescribing patterns in the three clusters. Despite the LPPN and PPNS subgroups having different initial severities of positive symptoms, and despite continued presence of positive symptoms at a higher level in only PPNS, clinicians seem to have elected to keep them at similar (though numerically lower) medication doses. This may have been due to relapse fears, or potentially the result of positive symptoms returning after short term dose reduction in the LPPN group; further work would be needed to decide this. It is also interesting to note the initial increase in CE in the LPPN group, followed by a decrease. This is likely due to the major persistent symptoms in this group being negative symptoms- clinicians may have increased doses early to treat positive symptoms, potentially tried decreasing them to attempt to reduce medication side effects which may have looked like negative symptoms, and then held doses steady given that they did not expect dose changes to further reduce negative symptoms.

#### Further information:

Chlorpromazine equivalent multiplication factors

| Generation | Trade name | Molecule | Chlopromazine equivalent |
|---|---|---|---|
| 2nd | Risperdal | Risperidone | 125 |
| 2nd | Risperdal Consta | Risperidone (injection) | 125 |
| 2nd | Zyprexa | Olanzapine | 33.33 |
| 2nd | Clozaril | Clozapine | 0.83 |
| 2nd | Seroquel | Quetiapine | 1.67 |
| 1st | Largactil | Chlorpromazine | 1 |
| 1st | Haldol | Haloperidol | 62.5 |
| 2nd | Invega | Paliperidone | 83.33 |
| 2nd | Invega Sustenna | Paliperidone palmitate (injection) | 83.33 |
| 2nd | Zeldox | Ziprasidone | 6.25 |
| 1st | Clopixol | Zuclopenthixol | 12 |
| 1st | Loxapac | Loxapine | 10 |
| 1st | Trilafon | Perphenazine | 20 |
| 2nd | Abilify | Aripiprazole | 25 |
| 1st | Nozinan | Methotrimeprazine | 2 |
| 2nd | Fluanxol | Flupenthixol | 60 |
| 1st | Modecate | Fluphenazine | 50 |

| | | | |
|---|---|---|---|
| 2nd | Saphris | Asenapine | 25 |
| 2nd | Latuda | Lurasidone | 6.25 |
| 2nd | | Iloperidone | 31.25 |
| 2nd | Serdolect | Sertindole | 20.83 |
| 1st | Stelazine | Trifluoperazine | 16.67 |
| 1st | Orap | Pimozide | 75 |

1. Pedregosa, F. *et al.* Scikit-learn: Machine Learning in Python. *J. Mach. Learn. Res.* **12**, 2825–2830 (2011).
